# Enhanced surveillance of monkeypox in Bas-Uélé, Democratic Republic of Congo: the limitations of symptom-based case definitions

**DOI:** 10.1101/2022.06.03.22275815

**Authors:** Gaspard Mande, Innocent Akonda, Anja De Weggheleire, Isabel Brosius, Laurens Liesenborghs, Emmanuel Bottieau, Noam Ross, Guy -Crispin Gembu, Robert Colebunders, Erik Verheyen, Ngonda Dauly, Herwig Leirs, Anne Laudisoit

## Abstract

**Background:** Following an outbreak of cases of vesicular-pustular rash with fever evocative of human monkeypox in Bas-Uélé province, Democratic Republic of Congo, surveillance was strengthened.

**Methods:** Households with at least one active generalized vesicular-pustular rash case were visited, and contact and clinical history information was collected from all household members. Whenever possible, skin lesion were screened by PCR for the monkeypox virus, followed by the varicella-zoster virus when negative for the former.

**Results:** PCR results were obtained for 77 suspect cases distributed in 138 households, of which 27.3% were positive for monkeypox, 58.4% for chickenpox, and 14.3% negative for both. Confirmed monkeypox cases presented more often with monomorphic skin lesions, on palms of hands, and on soles of feet. Integrating these three features into the case definition raised the specificity to 85%, but would miss 50% of true monkeypox cases. A predictive model fit on patient demographics and symptoms had 97% specificity and 80% sensitivity, but only 80% and 33% in predicting out-of-sample cases.

**Conclusion:** Few discriminating features were identified and the performance of clinical case definitions was suboptimal. Rapid field diagnostics are needed to optimize worldwide early detection and surveillance of monkeypox.

## Background

Monkeypox virus (MPXV) is a DNA virus belonging to the Poxviridae family, which includes vaccinia, cowpox and variola (smallpox) viruses. MPXV is currently the most prevalent human Orthopoxvirus and has been re-emerging since the eradication of smallpox in 1980. In humans, it causes a disease similar to smallpox, called monkeypox (Durski et al., 2018). Two clades of MPXV have been identified: the West African clade (MPXV-WA) occurring in forest areas situated west of Nigeria, and the Congo Basin clade (MPXV-CB) in Central Africa (Sklenovská and Van Ranst, 2018). After an incubation period of usually 7 to 14 days (range from 5 to 21 days), human monkeypox typically begins with systemic symptoms (i.e. fever, myalgia) followed by a rash, starting on the face before centrifugally spreading to other parts, including hands and feet. The skin lesions generally appear all at the same stage (monomorphic), evolving within 2 to 4 weeks from macules to papules, vesicles, pustules, crusts, and scabs. Complications are frequent and mortality may be up to 10% in children. People (primary cases) get infected through contact with infected animals during farming, hunting, or bushmeat preparation activities, and human-to-human transmission (secondary cases) also occurs by respiratory droplets or by close contact with infected lesions or bodily fluids (Diaz, 2021, Makhani et al., 2019, Petersen et al., 2019).

Human monkeypox was first described in 1970 in the Democratic Republic of the Congo (DRC) (Ladnyj et al., 1972). It is now found in more than half of the health zones of the country, reaching an incidence exceeding 5000 cases per year. More than 35 epidemics have been reported in other African countries and in recent years cases have also been increasing in returning travelers (Erez et al., 2019, Hobson et al., 2021, Mauldin et al., 2020, Rao et al., 2022, Yong et al., 2020) culminating in 2022 in an unprecedented multi-country outbreak (WHO, 2022). Since 2002, monkeypox is a notifiable disease in DRC and part of the nationwide implemented Integrated Disease Surveillance and Response (IDSR) system (DRC, 2011, 2012). All suspected clinical cases (and deaths) that meet the clinical case definition have to be reported weekly by each health zone to the national level (Hoff et al., 2017). As shown in Table 1, two case definitions of “suspect monkeypox cases” are provided in the national IDSR guideline: one for use in health facilities by nurses/doctors and a second one much broader (“fever and cutaneous rash”) designed for surveillance by community health workers.

**Table 1.** Clinical case definitions for monkeypox applied in the Democratic Republic of the Congo surveillance system and in this study.

|  |  |
| --- | --- |
| **DRC clinical case definition for use in health facilities (clinical case definition A in Osadebe et al., 2017)** | Any person presenting with sudden onset of high fever followed after a few days by a vesicular-pustular rash predominantly on the face, palms of the hands, and soles of the feet, or presence of at least 5 variola-like scars. |
| **DRC clinical case definition for community surveillance** | Any person presenting with fever and a cutaneous rash |
| **Clinical case definition as applied in our study** | Any person with an active generalized vesicular-pustular rash (i.e. presenting at least five cutaneous lesions such as macules, papules, vesicles, pustules, or crusts) |
| **Clinical case definition B (as evaluated in Osadebe et al., 2017)** | Any person with a rash with deep-seated/firm lesions AND either febrile prodrome or lymphadenopathy or lesion of palms or soles |

Due to the remoteness of the endemic areas confirmatory testing by PCR, only available in the national reference laboratory (“Institut National de Recherche Biomédicale”, INRB, Kinshasa), is rarely performed. As a result, the robustness of field surveillance based on clinical suspicion has so far been difficult to evaluate. Several diseases with generalized skin eruption may mimic monkeypox, such as chickenpox, measles, molluscum contagiosum, or rickettsioses. The field differential diagnosis between monkeypox and chickenpox is particularly challenging (Jezek et al., 1988, Leung et al., 2019, MacNeil et al., 2009) and coinfections are frequent (Hughes et al., 2020). Recent attempts to design clinical case definitions with a higher specificity for surveillance or care purposes have not yet undergone external validation (Osadebe et al., 2017). Following alerts from several health zones of the Bas-Uélé province, northeastern DRC (Aketi, Buta, and Titule; Figure 1) of a sharp increase of suspect monkeypox cases a first exploratory mission was conducted in 2016 and confirmed an ongoing outbreak of febrile illness with skin eruption, suspect monkeypox. It also identified important challenges and gaps impacting the quality of the surveillance and outbreak data, including the non-systematic registration of cases and the scarcity of sampling kits in the field. An active surveillance component was added, and access to confirmatory testing was reinforced through the project entitled “Strengthening academic capacity to respond to Monkeypox epidemics: discrimination and origin of eruptive fevers in the Democratic Republic of Congo (DRC)” (Laudisoit, 2017).

**Figure 1:**
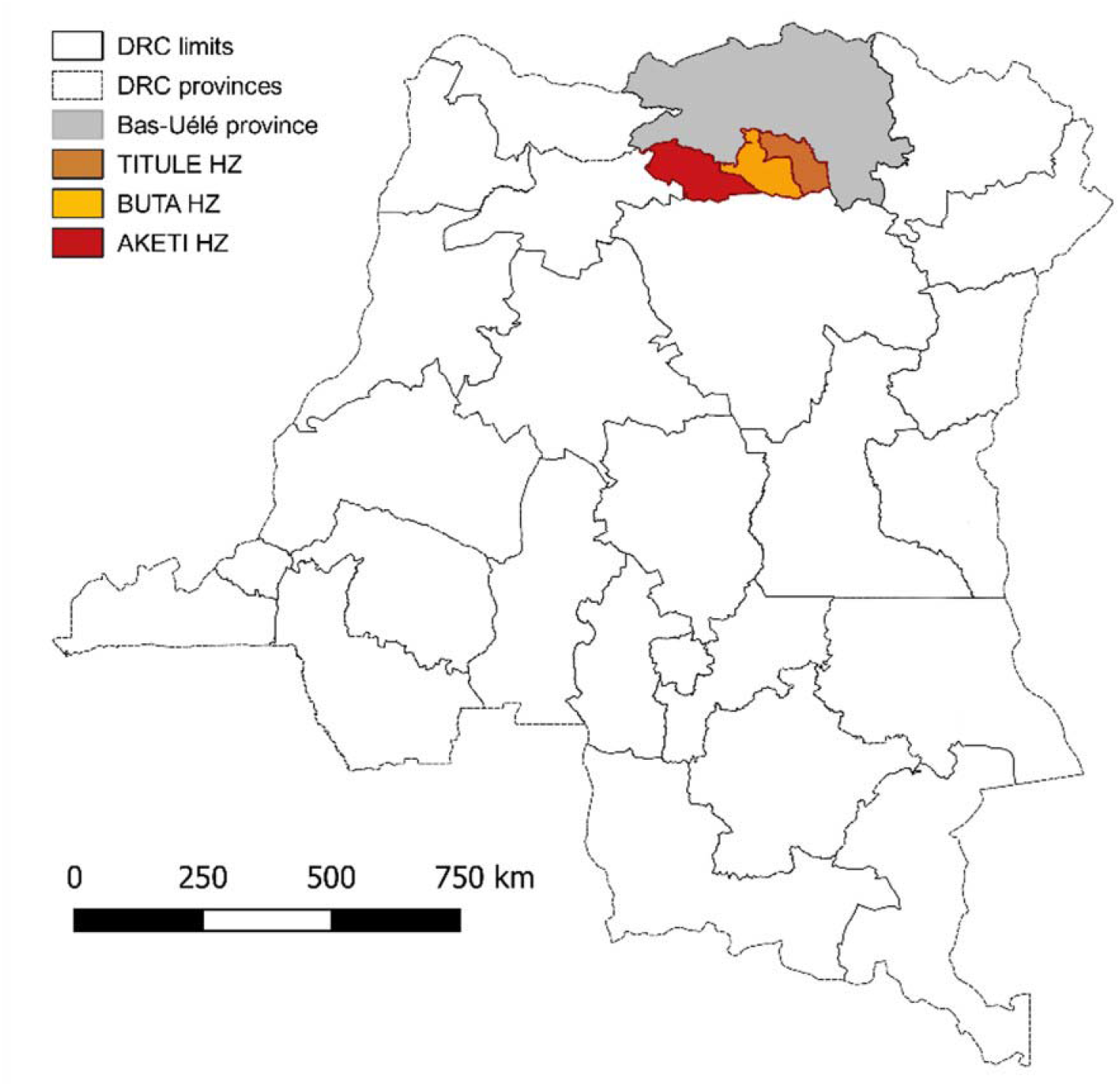
Map of the Democratic Republic of the Congo (DRC) indicating the Bas-Uélé province and the health zones of Aketi, Buta, and Titule (study area).

## Methods

### Study design, setting and population

In the framework of the project, enhanced active surveillance and household investigation of suspect monkeypox cases were first initiated in three health zones (Aketi, Buta, Titule) of the Bas-Uélé province, and later focused on the axis going North from Aketi (Figure 1), where most cases had been initially detected. This region is located in the lowland tropical rainforest of the Congo Basin, with a long rainy season from April to November, where the population lives mainly from subsistence farming, hunting, and small-scale livestock breeding.

The study population consisted of all suspect monkeypox cases as well as all members of their households, residing in locations accessible to trained study collaborators. For this study, a suspect monkeypox case was defined as “any person with an active generalized vesicular-pustular rash, i.e., presenting with five or more cutaneous lesions such as macules, papules, vesicles, pustules, or crusts, (Table 1)”, based on the DRC case definition for community surveillance (DRC, 2012). This study case definition was a bit stricter than that for community surveillance (a minimal number of lesions was required) but still aimed to largely capture any patients with active skin eruption that could be sampled for diagnostic evaluation.

### Enhanced surveillance and household investigation

Households in which at least one case of febrile eruptive skin disease was reported by community health care workers (RECO) were visited by the study team. The household visits were done during three field sessions (08-21/09/2017, 19-29/03/2018, 27/10-9/11/2018) and during the interval periods by trained registered nurses from the study areas.

At the household level, after having obtained written informed consent for household inclusion from the household head, epidemiological and clinical data were collected (number of household members, gender and age of each member, health zone/area and village of residence, geographical coordinates, past episodes of fever with skin eruption and smallpox vaccination status of each household member). In addition, for each individual considered as a suspect monkeypox case, signs and symptoms (focused on monkeypox features such as fever prodrome, type/stage of rash, ocular lesions, cervical lymphadenopathy), and risk factors (contact with sick person, animal contact, type of contact, time since contact) were collected using standardized questionnaires. In case of additional individual consent (or ascent by child plus consent of parent/tutor in case of children) and if sampling material was available, up to a maximum of 2 skin biopsies (crusts, pustules, or vesicle liquid) were collected following a strict standard operational procedure. One skin biopsy was stored ‘dry’ at room temperature in individual vials as per DRC monkeypox case management and surveillance guidelines and sent to INRB in Kinshasa (DRC, 2012). The second biopsy was stored in ethanol (70%) as a back-up and stored at UNIKIS. Venous blood was not collected. Data on outcomes or onward human-to-human transmission were not available, as cases and their households were not followed up longitudinally after the initial visit. During each field visit, information obtained in one household could lead to further investigational visits in other households of the same village (snowball enrolment).

### Laboratory procedures

Skin lesion samples were screened at the virology laboratory of INRB, Kinshasa, for Orthopoxviruses (OPXV) using an Orthopoxvirus-specific real-time in-house PCR assay (Osadebe et al., 2017). If the lesions tested negative for OXPV, a second real-time PCR assay targeting the varicella-zoster virus (VZV) was performed. OPXV positive samples were not tested further for VZV and were considered MPXV as no other circulating human pathogenic Orthopoxvirus causes a similar smallpox-like rash. Results were officially reported back from the reference laboratory (INRB) to the provincial central health bureau as positive, negative, or “neither MPXV nor VZV”, after interpretation of cycle threshold values.

### Evaluation of clinical case definitions

After this study was designed and approved, another research group published the results of a large case series of confirmed monkeypox and chickenpox cases and evaluated the DRC health facility case definition (called case definition A in this manuscript) and an alternative one (case definition B, elaborated to better discriminate both conditions) (Table 1). In addition, based on a receiver operating characteristic model including a set of 12 signs and symptoms, other combinations were proposed, since the performance of both case definitions A and B was unsatisfactory (Osadebe et al., 2017). We used our dataset retrospectively to also evaluate the diagnostic accuracy of both case definitions (with some slight adaptations) and other combinations, using the actual features that had been captured during our field study.

### Statistical analysis

Descriptive statistics, using medians and interquartile ranges for continuous variables and percentages for categorical variables, were used to summarize the results of the household investigations. Comparisons were calculated by Pearson’s chi-squared or Fisher exact test for categorical variables and the Kruskal Wallis test for continuous variables. Differences were considered statistically significant if p < 0.05. Diagnostic value and performance of signs/symptoms and different case definitions were expressed in sensitivity, specificity, positive and negative predictive value (PPV and NPV), and positive and negative likelihood ratios (LR + and LR-), using standard formulas. All statistical analyses were performed using Stata 15.0 software (StataCorp LP, College Station, TX, USA).

### Predictive model

We fit a model to predict PCR-confirmed cases using patient sex and age group, prior smallpox vaccination history, and presence of signs/symptoms. We generated interaction variables for every binary combination of variables, dropping interactions that were fully determined by other variables. We then fit a Bayesian logistic model to random training subset of 75% of PCR-confirmed cases, using a regularizing prior (Student’s t) on variable coefficient to optimize for out-of-sample prediction. We calculated the posterior odds ratios associated with each variable and interaction, and measured out-of-sample performance on the remaining 25% of cases. Model-fitting was performed in R 4.2.0 (RCoreTeam, 2022), using the Stan modeling framework (StanDevelopmentTeam, 2022) and brms package (Bürkner, 2017).

## Data Availability Statement

De-identified data and R code for the predictive model are deposited on Zenodo at https://dx.doi.org/10.5281/zenodo.6574450 (Mande et al. 2022).

## Results

Generalized skin eruption with fever evocative of human monkeypox was notified in 143 households, making a total of 948 people (median number of members/household: 6; interquartile range [IQR] 4-9) and reported by the RECO to the local health staff. These households were visited by the study teams between September 2017 and May 2019 (94 in the first year and 49 in the second year of the project). In a total of 106 households (106/143; 74.1%), representing 678 household members, the study team confirmed that at least one case responded to the study definition of suspected MPX. Overall, 138 patients (of whom 74 or, 54%, were aged <15 years) fulfilled the study definition of suspect monkeypox case (Table 2). The cases resided in 32 villages distributed in 12 health areas within three health zones (Aketi, Buta, Titule). Eighty-nine households (89/106; 84.0%) had one suspect case, 10 had two, four had three, one had four, and within two families, all members (6 and 7, respectively) had an active generalized vesicular-pustular rash fitting the study definition at the time of the household visit. During site visits, among households with active cases of skin eruption, 23/100 (n=100, info missing for 6 households) also reported recent and distant past episodes of fever with generalized skin eruption in their households (distant, i.e., >3 months earlier for 13; recent, i.e., within the last 3 months for 10).

**Table 2.**
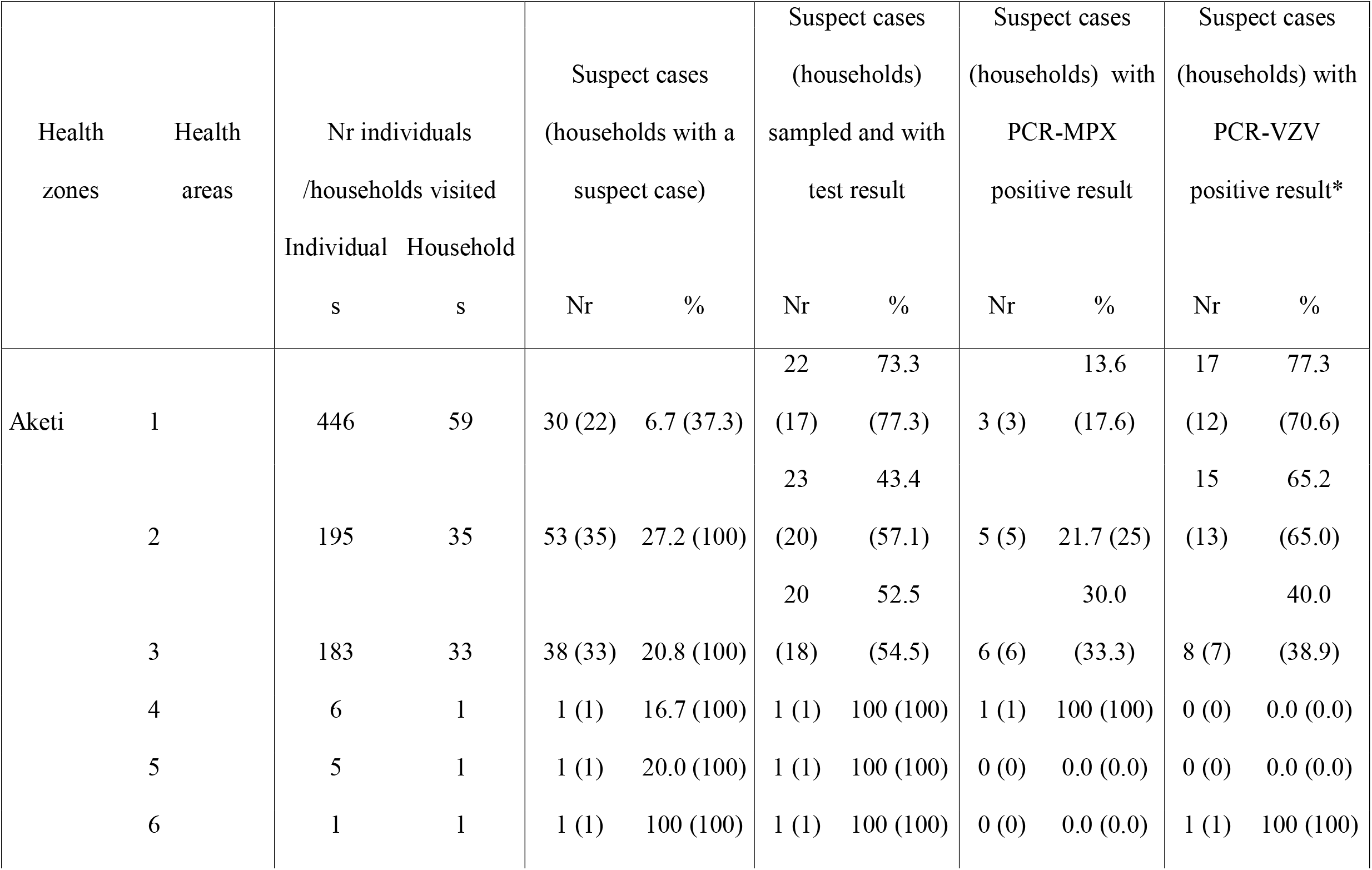
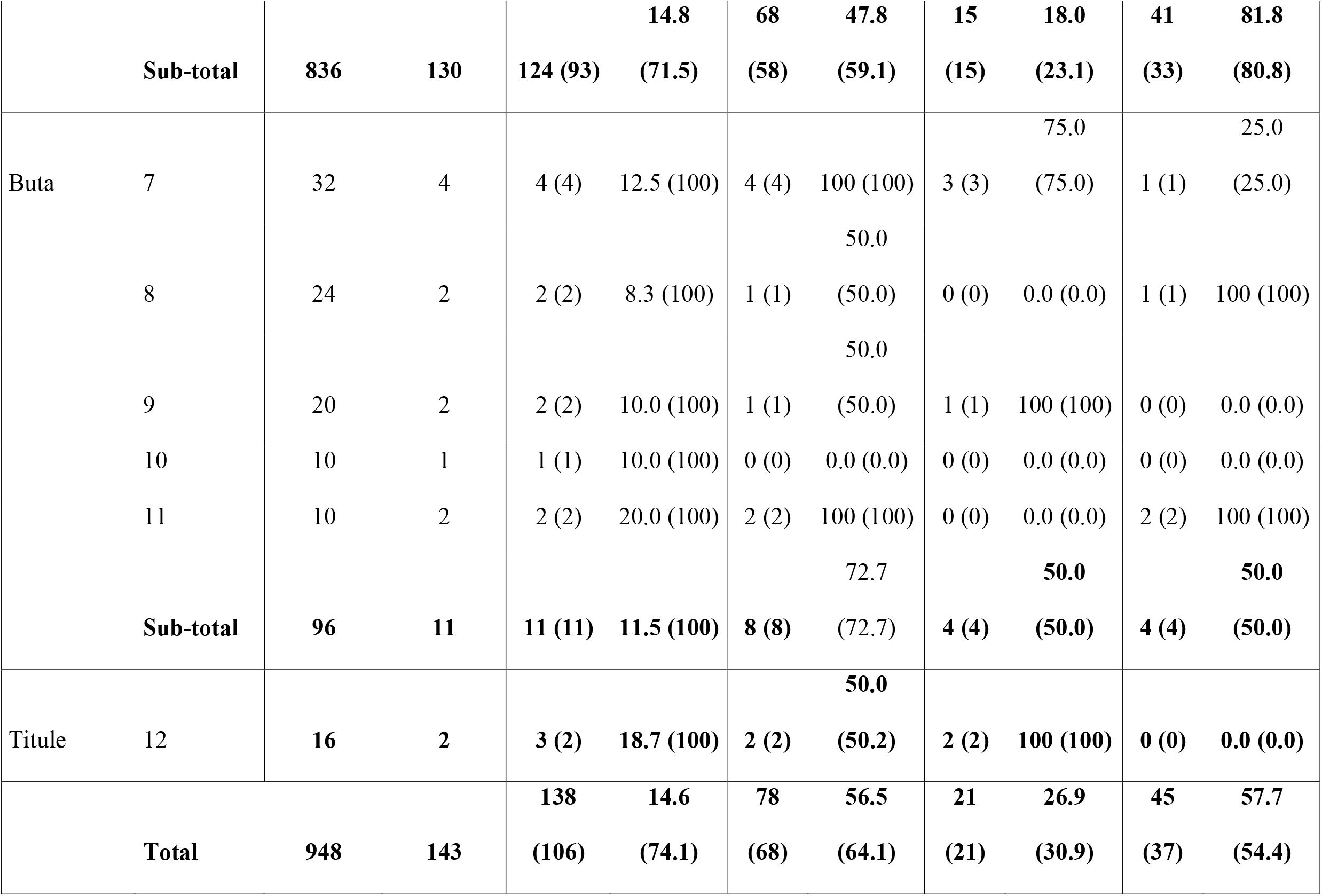
Overview of enhanced surveillance results from September 2017 to May 2019

Complete PCR results were obtained for 77 out of the 138 active suspect monkeypox cases (55.8%). Twenty-one of 77 patients (27.3%) tested positive for MPXV, and 45, out of the 56 MPXV-negatives, were positive for VZV. The remaining 11 cases remained negative for both MPXV and VZV. One suspect monkeypox case, negative for MPXV, was not tested for VZV. Next, we compared the socio-demographic, clinical characteristics, and risk factors between the 21 confirmed monkeypox cases, the 45 confirmed chickenpox cases, and the 11 cases reported as negative for both PCR assays (Table 3).

**Table 3.**
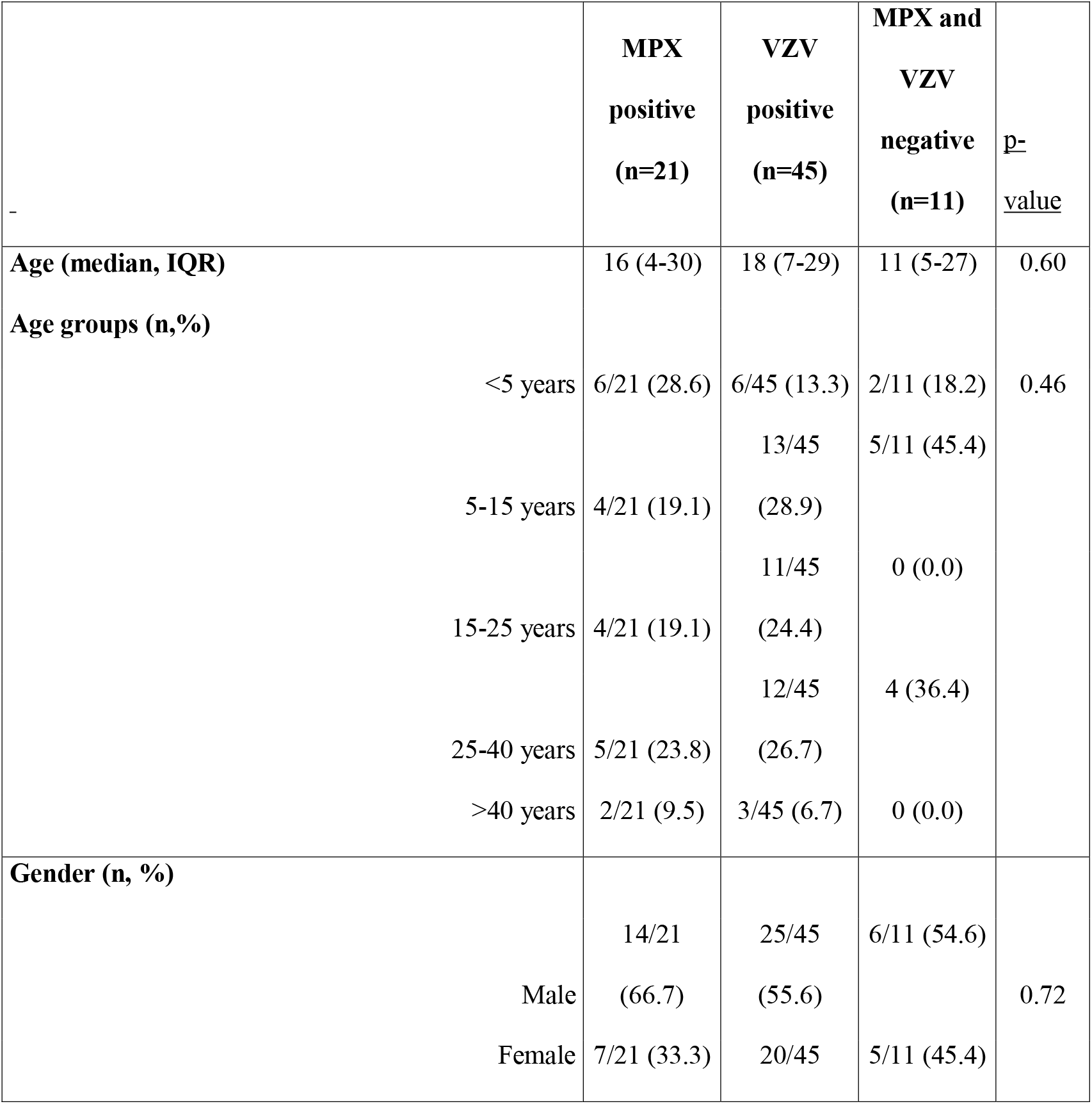
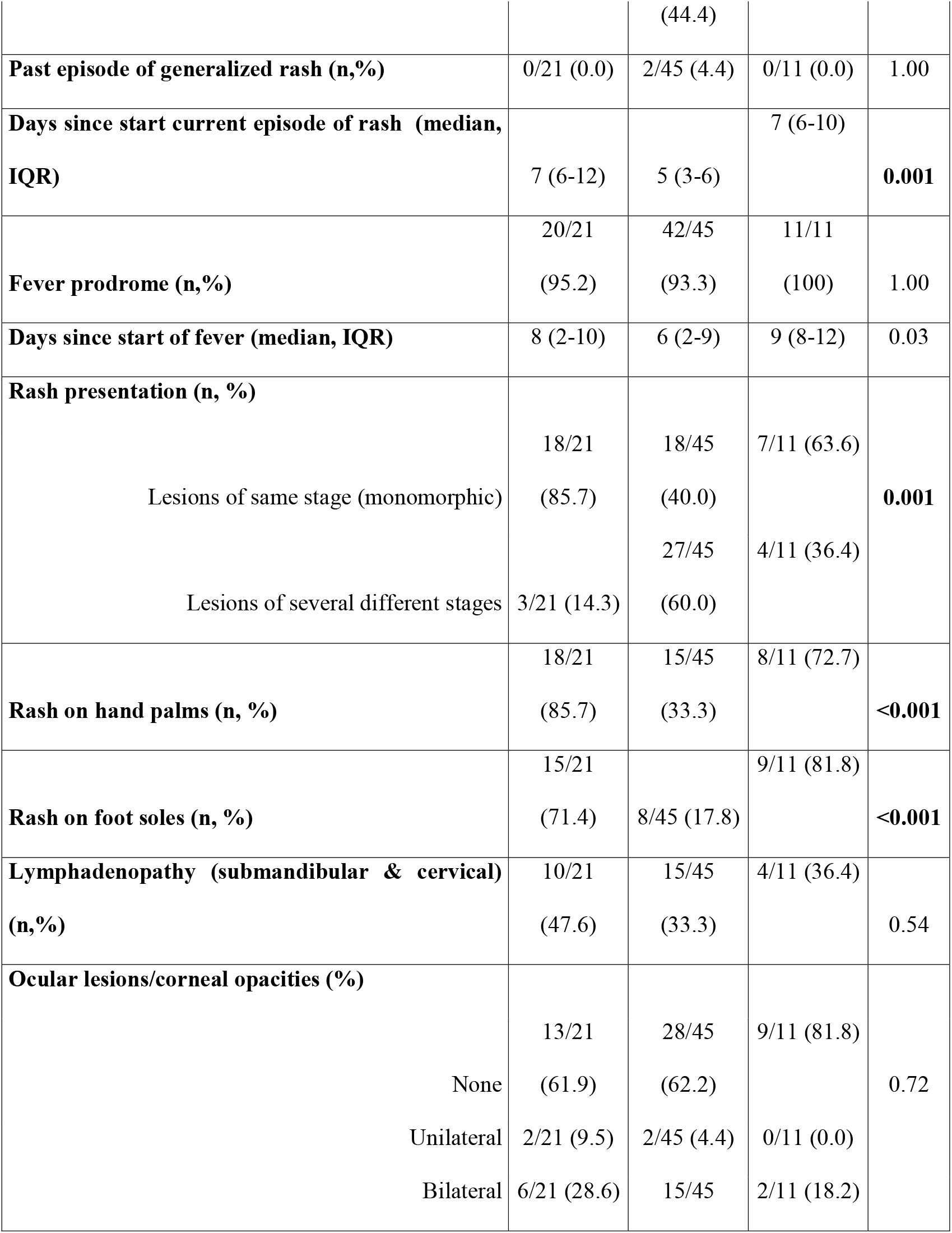
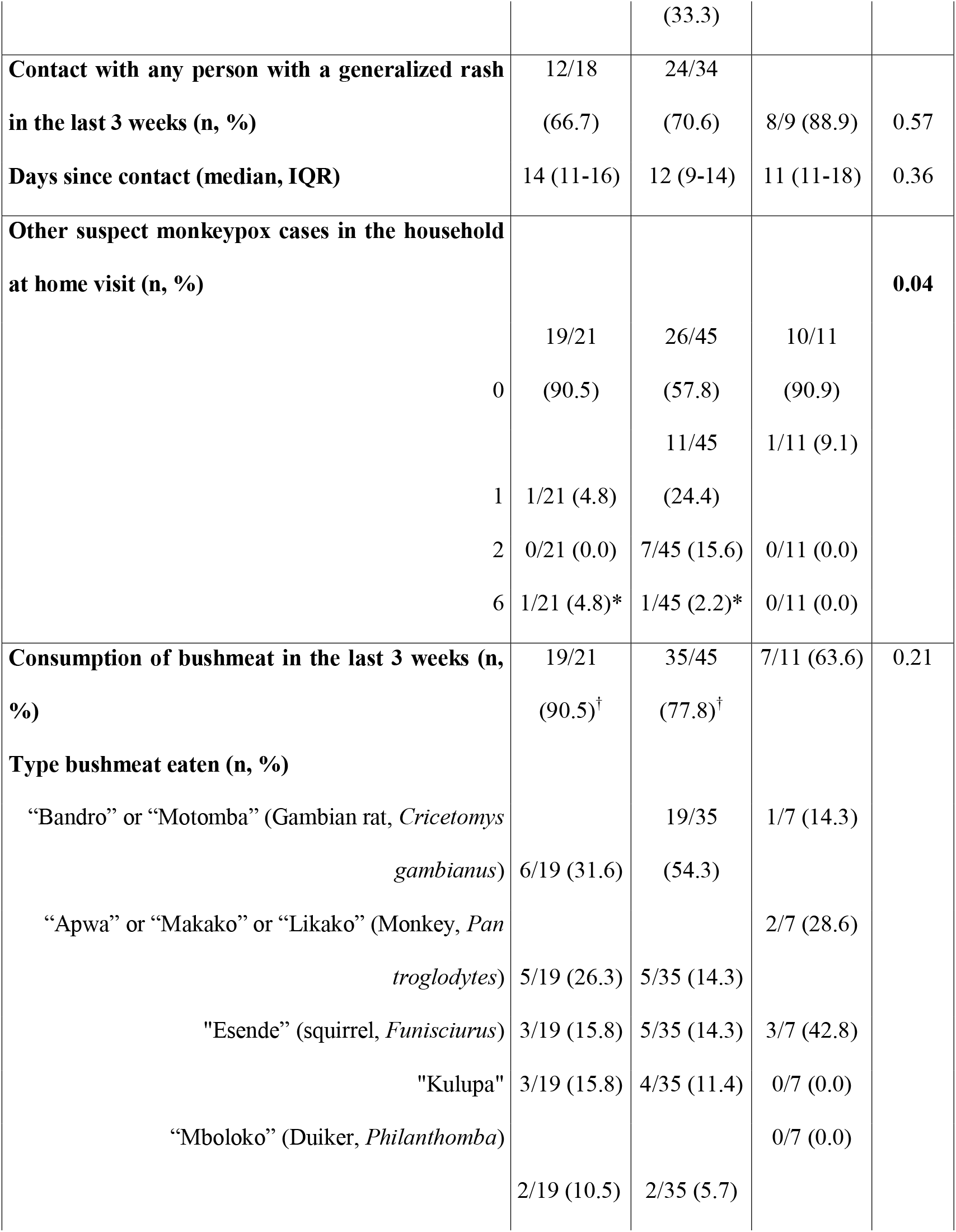
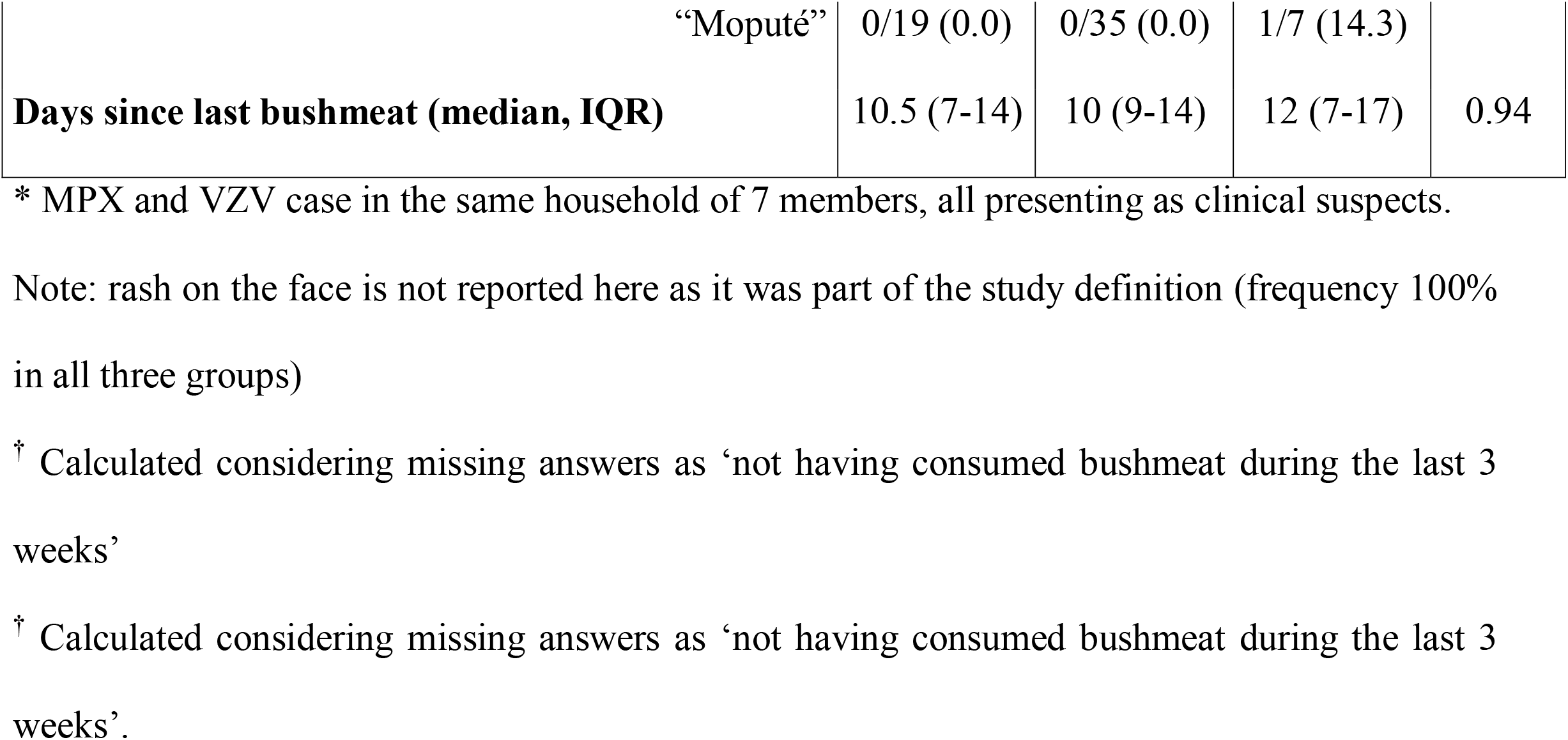
Socio-demographics, clinical characteristics, and risk factors for the confirmed monkeypox (MPXV positive), chickenpox (VZV positive), and PCR negative cases * MPX and VZV case in the same household of 7 members, all presenting as clinical suspects. Note: rash on the face is not reported here as it was part of the study definition (frequency 100% in all three groups).

Sociodemographic characteristics (age, gender) were similar in the three groups. In terms of clinical signs and symptoms, rash characteristics were the only symptoms that differed significantly between monkeypox and chickenpox cases. Monkeypox confirmed cases more often presented with cutaneous lesions in the same stage of the skin eruption (monomorphic) (85.7% vs 40.0% for the chickenpox cases; p=0.001); and more often had lesions on hands palms (85.7% vs 33.3%; p<0.001) and feet soles (71.4% vs 17.8%; p<0.001) compared to chickenpox patients. Forty percent of both confirmed monkeypox and chickenpox patients presented with ocular lesions/corneal opacities. Similarly, the frequency of cervical/submandibular lymphadenopathy was not significantly different between the two groups.

Contact with a person with generalized skin eruption within the past 3 weeks was reported in 70% of the cases of all three groups with no statistical difference. The presence of another suspect monkeypox case (according to the study definition) in the household tended to be more frequent in confirmed chickenpox cases than in MPX cases. Recent bushmeat consumption (giant pouched rat, primates, squirrels,…) was very common (>80%) in all three groups, with no statistical difference.

A similar number of confirmed monkeypox patients was reported from the three health areas (Area 1: 3/20, Area 2: 5/20, Area 3: 6/14) surveyed within the Aketi health zone. Of note, in one study household, MPXV- and VZV-positive cases were found concomitantly. Of note, in the 7-member household of which each individual presented with a rash, two persons were sampled, and one tested positive for MPX and the other patient tested positive for VZV. For the child, concurrent infection with MPX and VZV was not excluded. When comparing the clinical and socio-demographic features of suspect monkeypox cases with and without molecular results (Supplementary Table 1), no difference was observed except for the duration of fever (7 days [IQR 3-10] versus 9 days [6-12], respectively; p=0.008).

Based on the PCR results, the case definition used in this study led to a false positive rate of 73% (56 out of 77 tested samples were not confirmed as monkeypox). As detailed in Table 4, the case definition for “health facility” of the DRC national program would have decreased the false positive rate to 20% (corresponding to a specificity of 80%) but would have missed one-third of the true monkeypox cases (sensitivity of 67%). The proposed alternative case definition (18), would have an excellent sensitivity but a very low specificity (with >95% of false positives for monkeypox). All three identified discriminative features (“monomorphic rash”, “rash on palms”, “rash on soles”) had a weak confirming power for monkeypox (LR+ <3) when taken separately. Only the presence of all three features together has a good predictive value (LR+ >3), slightly better than that of the “health facility” case definition, increasing the probability of confirmed monkeypox above 50% (from a baseline frequency of 27% in suspect cases). In contrast, the absence of all three discriminative features would virtually exclude the diagnosis of monkeypox. The predictive model (Figure 2). had 97% specificity and 80% sensitivity in predicting PCR-confirmed monkeypox for in-sample cases, but only 80% specificity and 33% sensitivity for out-of-sample cases. Only one feature – the interaction of “rash on palms” in male patients” was a consistent predictor in >95% of model posterior samples. Other features that were positive predictors in >80% of posterior samples were all interactions including “rash on palms”, “rash on soles” and “monomorphic rash”. Prior vaccination against smallpox confirmed by presence of vaccinia vaccine scar was the strongest predictor against PCR-confirmed monkeypox (in 93% of samples).

**Table 4.**
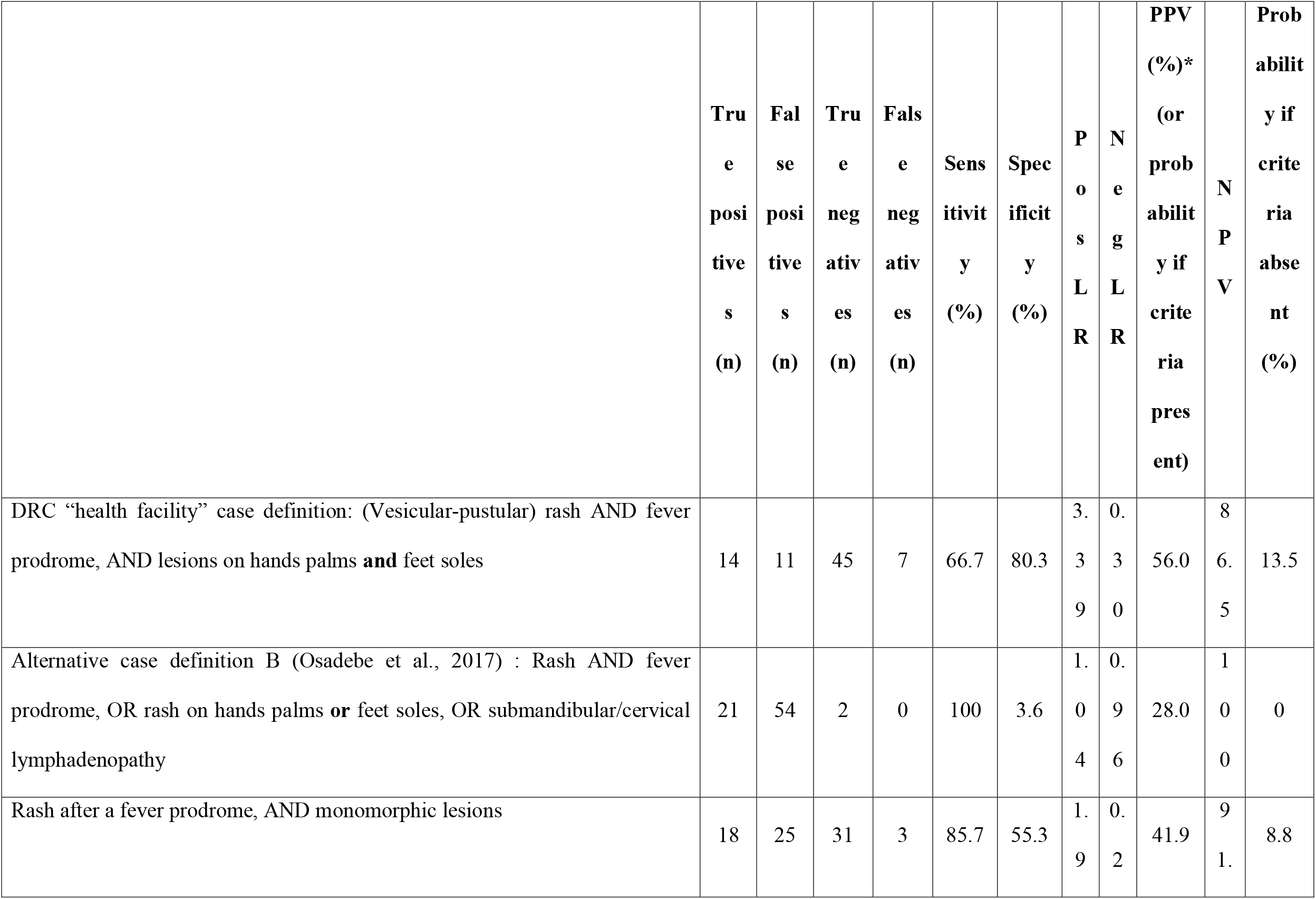
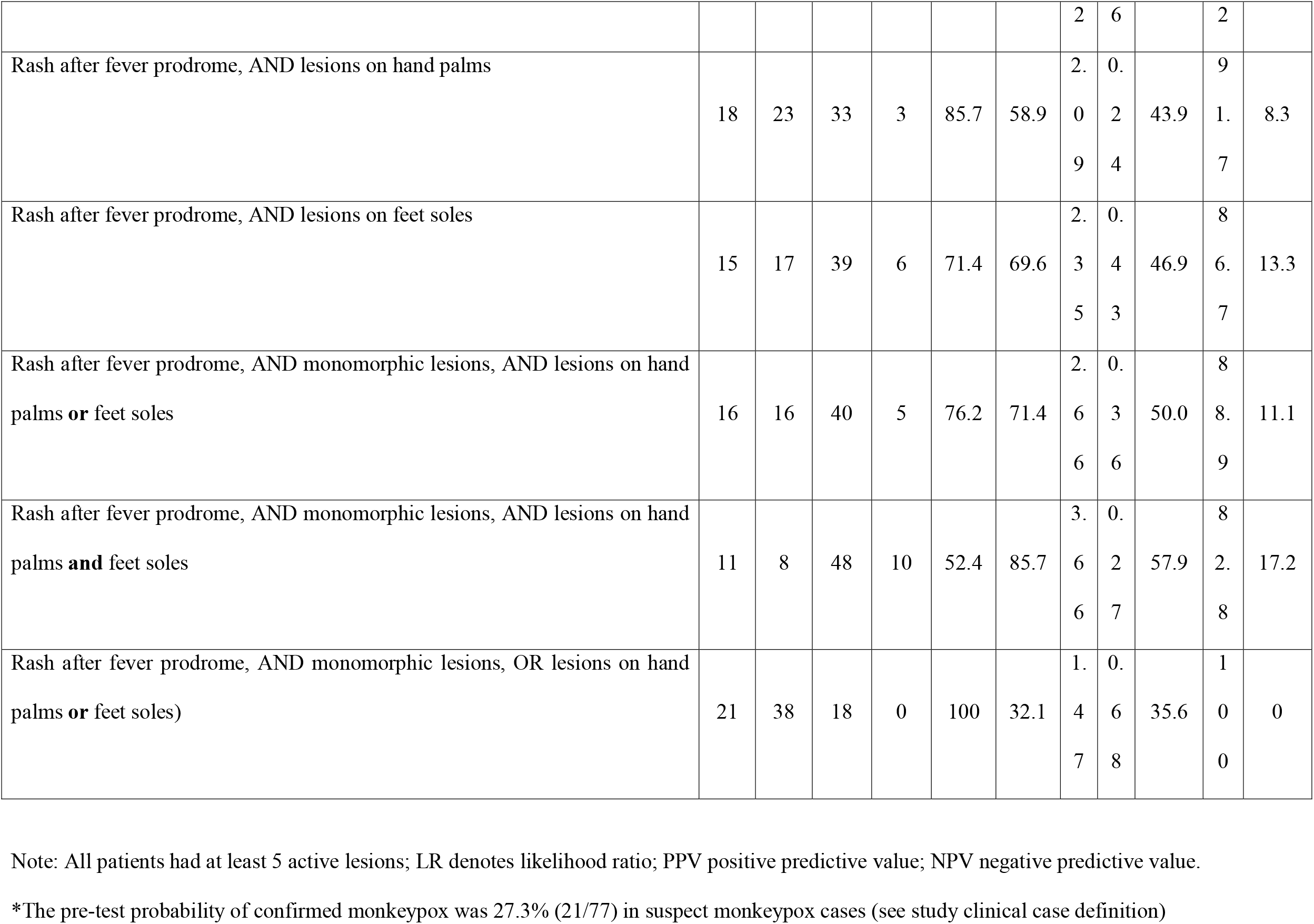
Diagnostic performance of “health facility” and alternative case definitions in the study dataset

**Figure 2.**
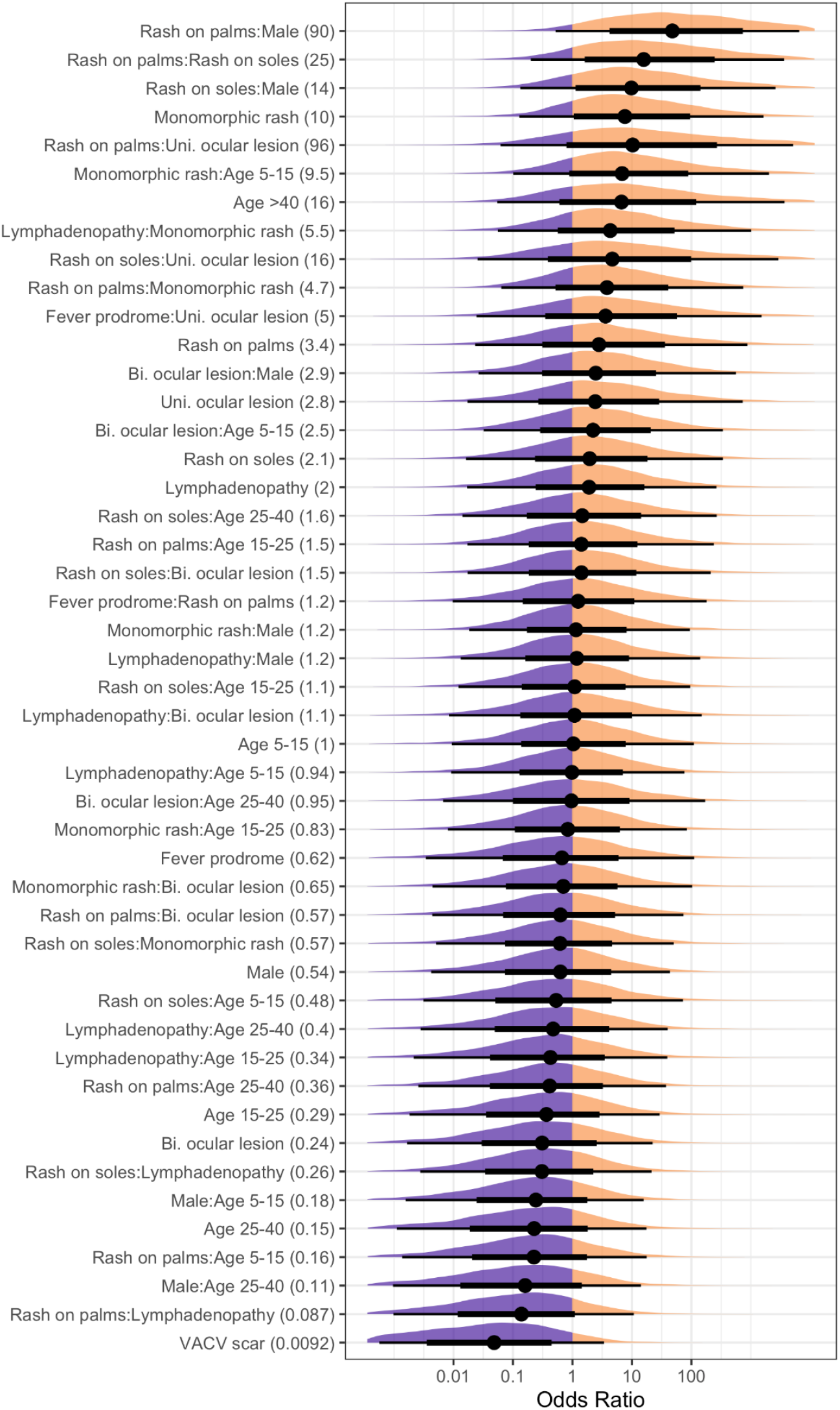
Coefficients for all variables and interactions (features) in the predictive model, scaled as odds ratios (OR). Mean OR is printed in the parentheses of each feature, with mean (dots), 66% posterior interval (thick lines), 95% posterior interval (thin lines), and full posterior shape (density plots) shown in the plot.

## Discussion

The enhanced surveillance of monkeypox in Bas-Uélé province by a dedicated “One Health” team confirmed that only 27% of suspect cases, as identified by an adapted “community” case definition, were confirmed as monkeypox. Most cases were finally diagnosed as chickenpox. The investigation did not identify clear epidemiological risk factors for monkeypox. Only three clinical features (monomorphic rash, rash on palms of hands, and rash on soles of feet) had some discriminative value for monkeypox, but no combination of symptoms and no alternative case definitions had a strong confirming power for decisive diagnosis.

This study has several limitations, inherent to the project and the health system in which it was embedded. First, the central objective of the project was education rather than research, with a progressive improvement of generic capacities towards monkeypox surveillance and detection. The clinical data collected during the project have thus evolved in terms of quality as the capacity increased over time, but some differences may have persisted between examinations made by medical doctors (during the field visits) and by nurses during the absence of supervision. Second, the intermittent provision of standard kits led to gaps in systematic skin sampling (which could be better addressed during the field visits). While we can consider that the subset of patients who were sampled for the molecular workup was selected at random, some bias may have occurred during these periods of higher availability. The technical difficulties related to sample preservation and transportation may have impacted the quality of the DNA and hence of the molecular results because laboratory staff and resources had on several occasions to be redirected to the reference diagnosis of Ebola during the successive outbreaks in DRC at that time. This has likely caused some false-negative results, as reflected by the clinical presentation of the 11 PCR-negative patients that was quite similar to that of confirmed monkeypox cases. Finally, as a result of the sequential INRB testing protocol, we cannot report the proportion of monkeypox cases concomitantly infected with VZV.

The low specificity of the case definition for community surveillance of monkeypox implies the inclusion of many false positives in both the epidemiological and clinical records scattered in peripheral health structures, with some under- and over-estimation of the actual caseload. Most captured cases were in fact chickenpox patients, confirming that the epidemiological characterization and clinical identification of monkeypox in Bas-Uélé are also seriously blurred by the co-endemicity of both conditions that share many similar and overlapping features (Jezek et al., 1988, Leung et al., 2019, MacNeil et al., 2009). It is expected that cases first captured by the very sensitive “community” case definition are assessed through the more restrictive “health facility” case definition, but this two-step clinical strategy may be not very well apprehended by less trained health workers. In addition, the latter case definition still includes a substantial number of non-monkeypox cases while it may create difficulties to classify the subgroup captured by the “community” definition which is secondarily rejected. Based on our limited dataset, we confirmed the discriminative value of some features such as the monomorphic aspect of the rash (all body lesions presenting at the same stage) and the preferential localization on palms and soles but also highlighted that their confirming power for monkeypox is limited. In contrast with others, we did not observe any predictive value for other characteristics such as cervical lymphadenopathy, ocular opacities, presence of other suspect cases in households - indicative of chickenpox in our dataset - or consumption of bushmeat, since they were all frequently reported in monkeypox and chickenpox confirmed patients. The negative correlation with prior vaccination against smallpox further supports high residual protection against monkeypox clinical diseases outcome, which is also supported by the generational divide in susceptibility since smallpox vaccination was discontinued (Rimoin et al., 2010). Looking for alternative case definitions, based on novel combinations of symptoms, did not substantially improve the diagnostic performance. Our predictive model also highlighted the importance of a monomorphic rash and localization on palms and soles. However, the model’s low predictive power, given its inclusion of all combinations of symptoms, shows the fundamental limitation on diagnosis without point-of-care tests. Because of its epidemic potential and substantial mortality and morbidity, accurate and swift diagnosis of monkeypox is crucial. There is a pressing need for peripheral laboratories with easy-to-use molecular diagnostic devices (Li et al., 2017) or the further provision and development of rapid point-of-care diagnostic tests such as ABICAP (Stern et al., 2016), suited to the remote harsh environments where monkeypox may break out at any moment.

Moreover, MPX antiviral treatment trials also should be conducted in Africa. For example, with the antiviral tecovirimat which received approval by both FDA and EMA for the treatment of MPX based on animal experiments (Grosenbach et al., 2018, Laudisoit et al., 2018). In addition, the efficacy of third generation smallpox vaccines in preventing MPX is being studied in the DRC. A smallpox-monkeypox vaccine has recently been validated by the FDA based on positive trials in other parts of DRC (Petersen et al., 2019).

In conclusion, this field investigation confirmed the difficulty observed elsewhere for health and community workers to clinically distinguish between monkeypox and chickenpox, two conditions that with similar symptoms but have increasingly divergent management. Combining lesion distribution and number, size, shape, and rugosity of lesions could potentially improve field discrimination between the two diseases. Development and field evaluation of breakthrough diagnostics and encouraging “One Health” intervention teams are key to timely and adequately address the variety and complexity of epidemic-prone diseases such as monkeypox, especially in areas with increasing evidence of co-circulation of zoonotic and anthroponotic pathogens (Bunge et al., 2022). The current outbreak of monkeypox outside Africa illustrates also the importance of intensified monkeypox surveillance in Africa among human and animal populations.

## Supporting information

Graphical abstract

## Data Availability

All de-identified data and R code for the predictive model are deposited on Zenodo at https://dx.doi.org/10.5281/zenodo.6574450
Citation : Mande, Gaspard, Akondab, Innocent, De Weggheleir, Anja, Brosius, Isabel, Liesenborghs, Laurens, Bottieau, Emmanuel, Ross, Noam, Gembu, Guy-Crispin, Colebunders, Colebunders, Verheyen, Erik, Dauly, Ngonda, Leirs, Herwig, & Laudisoit, Anne. (2022). Supplement to: Enhanced surveillance of monkeypox in Bas-Uele, Democratic Republic of Congo: the limitations of symptom-based case definitions. [Data set]. Zenodo. https://dx.doi.org/10.5281/zenodo.6574450

## Supplementary Material

The following supporting information can be downloaded at: Table S1: Comparison of sociodemographic and clinical characteristics of enrolled suspect monkeypox cases with versus without molecular results.

## Acknowledgments

The authors are grateful to the Bas-Uélé province authorities and in particular the Ministry of Health of Bas-Uélé province for administrative and logistical support. The researchers thank the main nurses of the visited health areas (Lizombe Réné and Egula Franck) and the biologists of the University of Kisangani (UNIKIS) and the Centre for Biodiversity Monitoring [Centre de Surveillance de la Biodiversité] for their help and scientific support during field surveys (Baelo Pascal, Malekani André, Musaba Prescott, Nebese Casimir, and Ngoy Steve), and National Institute of Biomedical Research (INRB) team in Kinshasa (Jean-Jacques Muyembe, Elisabeth Putuka and Steve Ahuka) for their scientific support. The authors are grateful to John Feigelson (Ecohealth Alliance) for producing the graphical abstract.

## Author Contributions

Conceptualization, Robert Colebunders, Erik Verheyen, Herwig Leirs and Anne Laudisoit; Data curation, Gaspard Mande, Anja De Weggheleire and Anne Laudisoit; Formal analysis, Anja De Weggheleire and Noam Ross; Funding acquisition, Robert Colebunders, Erik Verheyen, Herwig Leirs and Anne Laudisoit; Investigation, Gaspard Mande, Innocent Akonda, Emmanuel Bottieau, Guy-Crispin Gembu, Erik Verheyen and Anne Laudisoit; Methodology, Anja De Weggheleire, Emmanuel Bottieau, Anne Laudisoit and Noam Ross; Project administration, Robert Colebunders, Erik Verheyen and Anne Laudisoit; Supervision, Anja De Weggheleire, Emmanuel Bottieau, Erik Verheyen and Anne Laudisoit; Visualization, Anne Laudisoit and Noam Ross; Writing – original draft, Gaspard Mande, Innocent Akonda, Anja De Weggheleire and Emmanuel Bottieau; Writing – review & editing, All authors. All authors have read and agreed to the published version of the manuscript.

## Institutional Review Board Statement

The study was conducted following the Declaration of Helsinki and approved by the Ethics Committee of Antwerp University Hospital, Belgium (reference 18/01/012), and the Ethics Committee of the School of Public Health of the University of Kinshasa (approval number ESP/CE/077/2018). In addition to the household level consent signed by the household head for the home visit and data collection, individual written informed consent was obtained from each participant or its guardian in case of sample collection or picture capture.

