## Supplementary material for "Enhanced surveillance of monkeypox in Bas-Uélé, Democratic Republic of Congo: the limitations of symptom-based case definitions": Table S1

Table S1: Comparison of sociodemographic and clinical characteristics of enrolled suspect MPX cases with versus without molecular results.

|  | *Missing* | **MPX clinical suspect with lab result (77)** | *Missing* | **MPX clinical suspect without lab result (61)** | p-value |
| --- | --- | --- | --- | --- | --- |
| **Age (median, IQR)** | 0 | 17 (7-29) | 0 | 12 (7-27) | 0.54 |
| **Age groups (n,%)** |  |  |  |  |  |
| <5 years |  | 14 (18.2) |  | 13 (21.3) | 0.14 |
| 5-15 years |  | 22 (28.6) |  | 25 (41.0) |  |
| 15-25 years |  | 15 (19.5) |  | 5 (8.2) |  |
| 25-40 years |  | 21 (27.3) |  | 11 (18.0) |  |
| >40 years |  | 5 (6.5) |  | 3 (11.5) |  |
| **Gender (n, %)** | 0 |  | 0 |  |  |
| Male |  | 45 (58.4) |  | 36 (59.0) | 0.95 |
| Female |  | 32 (41.6) |  | 25 (41.0) |  |
| **Past episode of generalized rash (n,%)** | 0 | 2 (0.0) | 0 | 5 (8.2) | 0.24 |
| **Days since start current episode of rash (median, IQR)** | 4 | 6 (4-9) | 2 | 7 (5-10) | 0.32 |
| **Fever prodrome (n,%)** | 0 | 73 (94.8) | 2 | 58 (98.3) | 0.39 |
| **Days since start of fever (median, IQR)** | 10 | 7 (3-10) | 4 | 9 (6-12) | **0.008** |
| **Rash presentation (n, %)** | 0 |  | 6 |  |  |
| Lesions of a single-stage |  | 43 (55.8) |  | 26 (47.3) | 0.33 |
| Lesions of several stages |  | 34 (44.2) |  | 29 (52.7) |  |
| **Rash on hand palms (n, %)** | 0 | 41 (53.3) | 3 | 33 (56.9) | 0.67 |
| **Rash on foot soles (n, %)** | 0 | 32 (41.6) | 3 | 21 (36.2) | 0.53 |
| **Lymphadenopathy** **(submandibular & cervical) (n,%)** | 0 | 29 (37.7) | 1 | 31 (51.7) | 0.10 |
| **Ocular lesions/ conjunctivitis (%)** | 0 |  | 0 |  |  |
| None |  | 50 (64.9) |  | 36 (59.0) | 0.65 |
| Unilateral |  | 4 (5.2) |  | 2 (3.3) |  |
| Bilateral |  | 23 (29.9) |  | 23 (37.7) |  |
