## Supplementary figures and images for "Enhanced surveillance of monkeypox in Bas-Uélé, Democratic Republic of Congo: the limitations of symptom-based case definitions"

### Graphical abstract

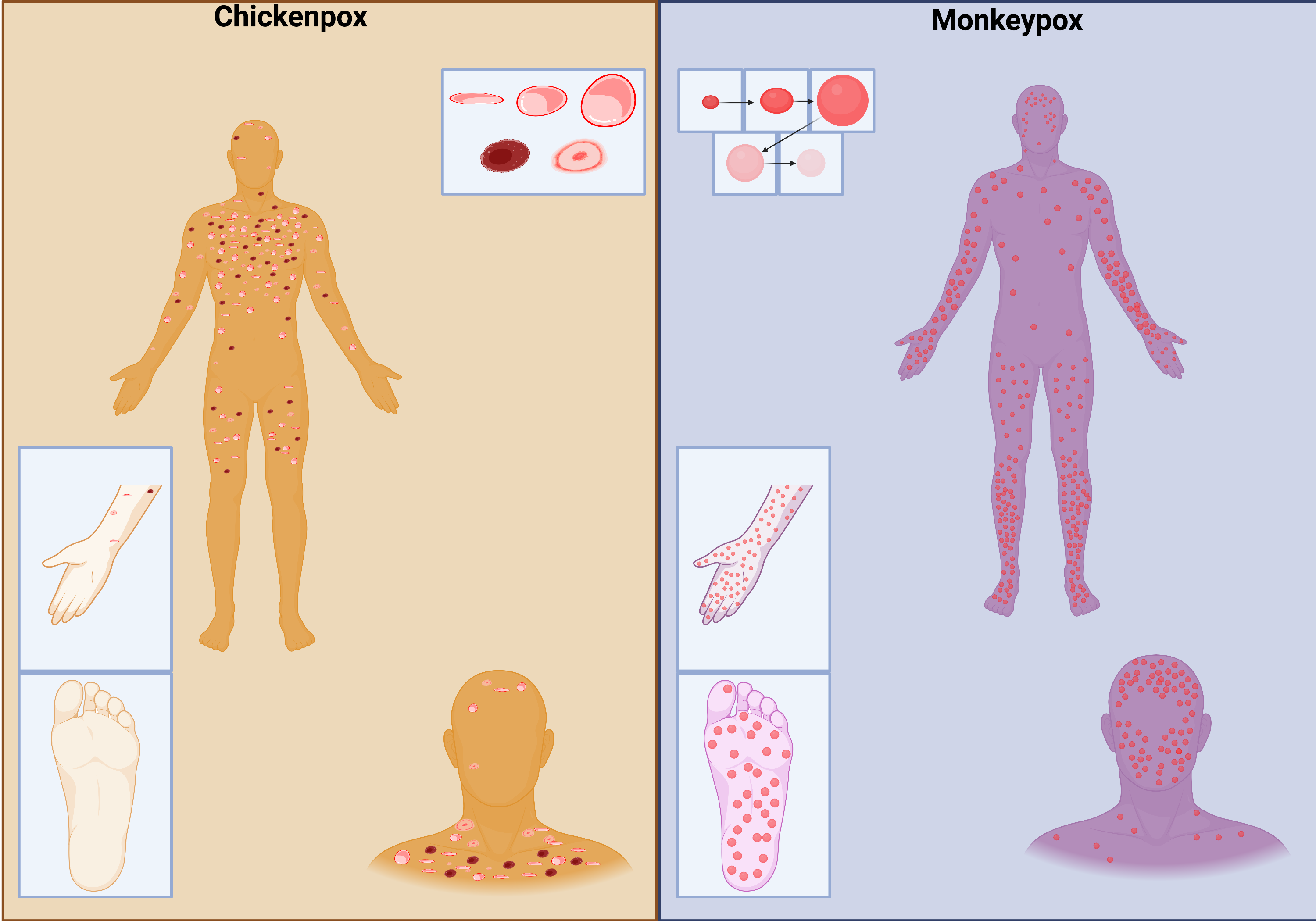
